# Safety Monitoring of Multiple Health Outcomes Following 2023–2024 COVID-19 Vaccination among Medicare Beneficiaries Aged 65 Years and Older in the United States

**DOI:** 10.1101/2025.01.03.25319975

**Authors:** Joann F. Gruber, Michelle Ondari, Carla E. Zelaya, Chunyi Xia, Fengdi Zhang, Jessica R. Hervol, Jin Ye, Meng Chen, Yutong Qin, Mao Hu, Yoganand Chillarige, Richard A. Forshee, Steven A. Anderson

**Affiliations:** U. S. Food & Drug Administration, 10903 New Hampshire Ave., Building 71, Silver Spring, MD 20993, United States; Acumen, LLC, 500 Airport Blvd. Suite 100, Burlingame, CA 94010, United States

**Keywords:** COVID-19 vaccine, safety, Pfizer-BioNTech, Moderna, Novavax, anaphylaxis

## Abstract

**Background:** COVID-19 vaccines are well-established as safe. However, continued surveillance of COVID-19 vaccines is important to ensure the safety of newly formulated vaccines. This study evaluated the association between vaccination with 2023-2024 formula COVID-19 vaccines and multiple health outcomes among Medicare beneficiaries aged 65 years and older in the United States.

**Methods:** The study used health plan data from the Medicare Fee-for-Service (FFS) claims database and extended from September 2023 to April 2024. We monitored the uptake of 2023–2024 COVID-19 vaccines (Pfizer-BioNTech, Moderna, Novavax), and where case counts were available, we used a self-controlled case series design to assess the association between vaccination and prespecified health outcomes. We used conditional Poisson regression to estimate incidence rate ratios (IRRs), attributable risks (ARs) and corresponding 99% confidence intervals (CIs). Analyses were adjusted for outcome seasonality, event-dependent observation time for outcomes with high case fatality rates, and outcome misclassification where feasible.

**Results:** Approximately 7.6 million Medicare FFS beneficiaries received a 2023–2024 COVID-19 vaccination. There was an even distribution of people who received Pfizer-BioNTech (3,689,356; 48.80%) or Moderna (3,841,245; 50.80%) vaccinations and very few who received Novavax (30,171; 0.40%). There was a statistically significant elevation in anaphylaxis risk associated with 2023–2024 Pfizer-BioNTech vaccines in the seasonality-adjusted analysis (IRR: [99% CI: 1.07, 15.30]) that was no longer statistically significant after accounting for potential outcome misclassification (IRR: 3.90 [99% CI: 0.49, 30.90]). Anaphylaxis cases attributable to 2023–2024 Pfizer-BioNTech vaccination were rare (AR per 100,000 doses: 0.09 [99% CI: −0.08, 0.25]). No other statistically significant elevations in risk were observed.

**Conclusion:** There were no new safety signals identified following 2023–2024 COVID-19 vaccinations among U.S. Medicare beneficiaries aged 65 years and older. A potential, but rare, elevation in anaphylaxis incidence rates following 2023–2024 Pfizer-BioNTech COVID-19 vaccination was observed.

**HIGHLIGHTS:**

- We studied 7,560,772 Medicare enrollees ≥65 years old who received 2023–2024 COVID-19 vaccines
- No new safety signals were identified in the Medicare population ≥65 years old
- A potential–but rare–elevation in anaphylaxis risk was associated with vaccination
- No increased risk of any other prespecified health outcomes was found

## 1 Introduction

The first coronavirus disease 2019 (COVID-19) vaccines were introduced in the United States (U.S.) in December 2020 to protect individuals from severe COVID-19 and related complications such as hospitalization and death [1]. Vaccination continues to be the most effective strategy for preventing severe COVID-19 and is particularly important for persons aged 65 years and older who are at higher risk of severe complications following COVID-19 [2–7].

The U.S. Food and Drug Administration (FDA) has monitored the safety of prior formulations of COVID-19 vaccines among persons aged 65 years and older, and no conclusive safety concerns have been identified for this age group [8, 9]. Self-controlled case series (SCCS) studies evaluating the risk of outcomes following COVID-19 vaccination of persons aged 65 years and older from December 2020 to May 2022 found no statistically significant increased risk of acute myocardial infarction, Bell’s palsy, disseminated intravascular coagulation, immune thrombocytopenia, or myocarditis/pericarditis, with inconclusive results for pulmonary embolism risk [8]. Consistent findings have been observed in international studies in similar age groups [10–12]. In addition, multiple studies conducted among persons aged 65 years and older found no clear or consistent evidence of an association between stroke risk and COVID-19 vaccination [13–19].

In September 2023, FDA approved the updated 2023–2024 formula COVID-19 mRNA vaccines Comirnaty (Pfizer-BioNTech) and Spikevax (Moderna) for persons 12 years and older and authorized the 2023–2024 formula vaccines under emergency use for persons 6 months through 11 years [20]. In October 2023, the 2023–2024 formula Novavax COVID-19 adjuvanted vaccine was authorized under emergency use for individuals 12 years and older [21]. The 2023– 2024 COVID-19 vaccines were updated to include a monovalent component to protect against the SARS-CoV-2 Omicron XBB.1.5 subvariant, expected to be the most prevalent strain during 2023–2024 [20, 21]. In September 2023, the Advisory Committee on Immunization Practices (ACIP) recommended all persons 6 months and older receive a 2023–2024 COVID-19 vaccine, and subsequently in February 2024, ACIP recommended that persons 65 years and older receive an additional dose [22, 23].

Post-marketing surveillance of COVID-19 vaccines continues to be important to ensure the safety of newly formulated vaccines recommended for widespread administration. The objective of this study was to assess the safety of the 2023–2024 COVID-19 vaccines administered to Medicare Fee-for-Service (FFS) beneficiaries aged 65 years and older. This manuscript summarizes observed incidence rates of multiple prespecified health outcomes following the administration of 2023–2024 COVID-19 vaccines and presents results from an SCCS study evaluating the association between these vaccinations and health outcomes, accounting for time-invariant confounders [24].

## 2 Methods

### 2.1 Study Overview

We monitored the uptake of the 2023–2024 COVID-19 vaccines among Medicare beneficiaries aged 65 years and older. Following vaccination, we assessed the incidence rates of 14 prespecified health outcomes that included serious health outcomes following other vaccinations or outcomes potentially related to novel platforms, adjuvants, or COVID-19 severity. These outcomes included acute myocardial infarction; anaphylaxis; appendicitis; deep vein thrombosis; disseminated intravascular coagulation, encephalitis or encephalomyelitis (including acute disseminated encephalomyelitis); Guillain-Barré syndrome (GBS); hemorrhagic stroke; a composite outcome of myocarditis, pericarditis, or co-occurring myocarditis and pericarditis (referred to as myocarditis/pericarditis); non-hemorrhagic stroke; non-hemorrhagic stroke or transient ischemic attack (TIA); pulmonary embolism; transverse myelitis; and thrombosis with thrombocytopenia syndrome.

We conducted power analyses to determine if there was sufficient power to conduct SCCS analyses, comparing the incidence rate of each outcome in risk intervals (which were defined as periods of hypothesized excess outcome risk following vaccination) and control intervals, which included all other time in the post-vaccination observation period not in the risk interval or washout period (Figure 1). We also examined the effect of concomitant vaccination in SCCS analyses for outcomes with an elevation in incidence following vaccination, meeting a prespecified p-value threshold.

**Figure 1.**
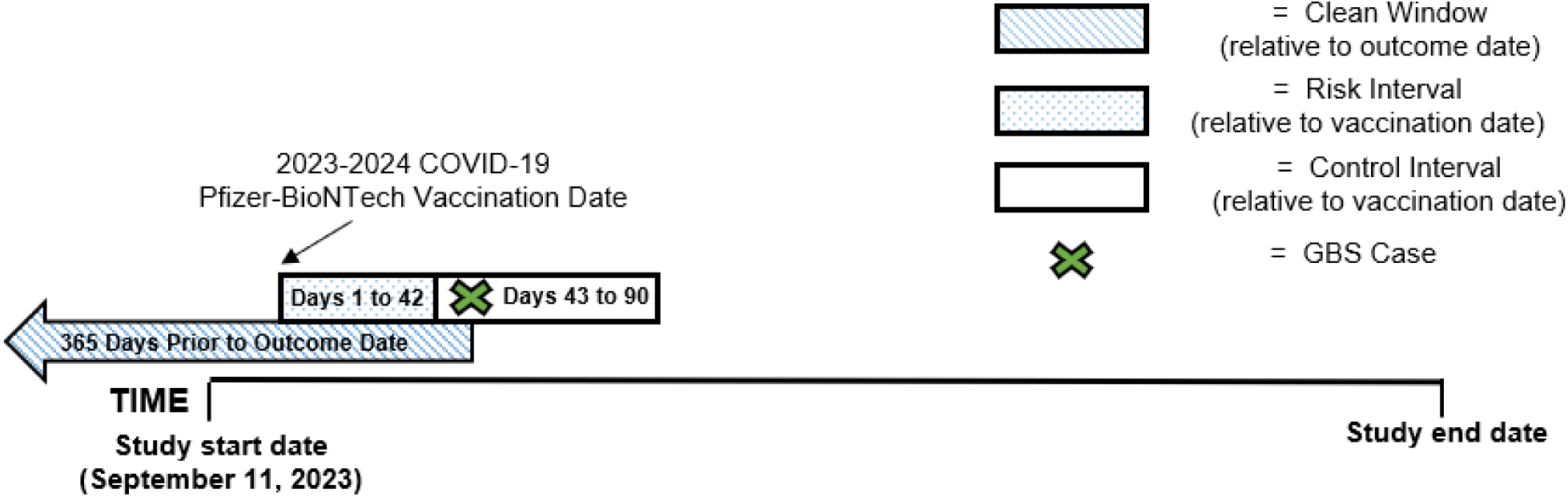
Example of study follow–up for the outcome of GBS following 2023 –2024 Pfizer–BioNTech vaccination.

### 2.2 Data Sources, Study Population, and Study Period

The study captured vaccination and longitudinal health information for Medicare FFS (Medicare Parts A and B, but not Part C) beneficiaries aged 65 years and older using claims data from the CMS Medicare Shared Systems Data. The Medicare Enrollment Database and Common Medicare Environment were used to capture individuals’ demographic characteristics and information on death. Information on nursing home residency status was captured using the Minimum Data Set 3.0.

Study participants were required to receive a 2023–2024 formula COVID-19 vaccine after the brand-specific approval or authorization date. They were also required to be enrolled in a Medicare FFS plan and be age 65 years or older at the time of vaccination. We excluded persons with an unspecified COVID-19 vaccine code on the date of their first observed vaccination, persons with codes for multiple COVID-19 vaccine brands on the same day or within 3 days of their first observed vaccination, and persons with missing age or sex information. For estimation of outcome incidence rates and SCCS analyses, persons were also required to have continuous medical enrollment in a Medicare FFS plan in the 365 days prior to their first observed COVID-19 vaccination; for SCCS analyses, persons were also required to have an incident outcome during the outcome-specific observation period (Figure 1).

The study period started on September 11, 2023, the earliest date of approval/authorization of any 2023–2024 COVID-19 vaccine, and extended to April 6, 2024.

### 2.3 Exposure

The exposure of interest was receipt of any of the approved or authorized 2023–2024 COVID-19 vaccines which were identified using Current Procedural Terminology (CPT)/Healthcare Common Procedure Coding System (HCPCS) codes, or National Drug Codes (NDCs) [25]. We restricted the analysis to persons with single doses of a known vaccine brand (2023–2024 Pfizer-BioNTech, Moderna, or Novavax) or multiple doses of the same vaccine brand (homologous doses) [26]. To ensure we were not capturing duplicate doses for the same COVID-19 vaccine administration, we deduplicated vaccination codes observed within 3 days of each other into a single administration, with the administration date set to the date of the first observed vaccination code. For the estimation of outcome incidence rates and SCCS analyses, we included COVID-19 doses up until a vaccine cutoff date that varied by outcome (range December 3, 2023 to March 21, 2024) to allow sufficient time to observe 90% outcome completeness in risk and control intervals following vaccination in claims data.

### 2.4 Outcomes and Follow-Up

Our study monitored 14 incident outcomes following COVID-19 vaccination. Outcomes were identified using International Classification of Disease, Tenth Revision, Clinical Modification (ICD-10-CM) codes [25]. To ensure we identified incident outcomes that were unrelated to previous disease manifestations, a corresponding outcome could not have occurred during a prespecified period (e.g., 365 days) prior to the observed outcome (i.e., clean window, Figure 1). The claims settings, risk and control intervals, and clean windows for each outcome are defined in Table 1.

**Table 1:** Administrative claims-based outcome definitions for Medicare Fee-For-Service beneficiaries aged ≥65 years.

| Outcome<sup>a</sup> | Claims Setting<sup>b</sup> | Clean Window (days) | Risk Interval (days) | Control Interval (days) |
| --- | --- | --- | --- | --- |
| Acute myocardial infarction | IP | 365 | 1–28[39] | 29–90 |
| Anaphylaxis | IP, OP–ED | 30 | 0–1[40] | 3–16 |
| Appendicitis | IP, OP–ED | 365 | 1–42[41] | 43–90 |
| Deep vein thrombosis | IP, OP/PB | 365 | 1–28[42,43] | 29–90 |
| Disseminated intravascular coagulation | IP, OP–ED | 365 | 1–28[44] | 29–90 |
| Encephalitis/encephalomyelitis | IP | 183 | 1–42[45] | 43–90 |
| Guillain–Barré syndrome | IP-primary position only | 365 | 1–42[46] | 43–90 |
| Hemorrhagic stroke<sup>c</sup> | IP | 365 | 1–21[47] & 22–42[47] | 43–90 |
| Myocarditis/Pericarditis<sup>d</sup> | IP, OP–ED &<br>IP, OP/PB | 365 | 1–7[48] | 8–90 |
| Non–hemorrhagic stroke (NHS)<sup>c</sup> | IP | 365 | 1–21[47] & 22–42[47] | 43–90 |
| NHS or Transient ischemic attack (TIA)<sup>c</sup> | IP (NHS & TIA),<br>OP–ED (TIA) | 365 | 1–21[47] & 22–42[47] | 43–90 |
| Pulmonary embolism | IP | 365 | 1–28[42,43] | 29–90 |
| Transverse myelitis | IP, OP–ED | 365 | 1–42[49] | 43–90 |
| Thrombosis with thrombocytopenia syndrome (TTS) | TTS episode<sup>e</sup> | 365 | 1–28[50] | 29–90 |

Acronyms: IP: inpatient, OP: outpatient, OP–ED: outpatient emergency department, PB: provider or professional services, NHS: non–hemorrhagic stroke, TIA: transient ischemic attack, TTS: thrombosis with thrombocytopenia syndrome
<sup>a</sup>The algorithms and code lists used to identify incident outcomes can be referenced in the study protocol.[25]
<sup>b</sup>IP was defined as inpatient facility claims. OP/PB was defined as outpatient facility claims or professional/provider claims that contain at least one non–laboratory place of service (independent laboratory place of service code = 81). OP–ED was defined as outpatient facility claims in emergency departments and is a subset of the OP/PB care setting.
<sup>c</sup>Outcome definitions includes two different risk intervals to assess the post–vaccination risk of outcomes.
<sup>d</sup>There are two myocarditis/pericarditis outcome definitions. One definition identifies outcomes in any setting (IP, OP/PB); the second identifies outcomes in the IP or OP–ED settings only.
<sup>e</sup>TTS was a combined outcome consisting of a thrombotic outcome (e.g., acute myocardial infarction, deep vein thrombosis, etc.) and a thrombocytopenia outcome (defined in the IP, OP/PB setting). The overall setting definition for each thrombotic outcome depends on individual setting definitions for each of the respective components.

Persons were followed for the length of the outcome-specific observation period, which was 90 days following each COVID-19 vaccine dose for all outcomes with the exception of anaphylaxis, which was 16 days. Follow-up was censored at the first occurrence of disenrollment, death, receipt of a heterologous COVID-19 vaccine dose, receipt of a homologous COVID-19 vaccine dose after the vaccine cutoff date, or study period end.

### 2.5 Covariates

Covariates, including demographic information, medical conditions, and concomitant vaccination status, were defined using claims data and used to describe the populations of interest. Demographic variables were defined at the time of COVID-19 vaccination using enrollment data. Medical conditions were defined using ICD-10-CM codes in any care setting in the 183 days prior to the first COVID-19 vaccine dose. Concomitant vaccination was defined as receipt of influenza, respiratory syncytial virus (RSV), pneumococcal conjugate, or zoster vaccines, defined using HCPCS/CPT and NDCs, on the same date as the COVID-19 vaccination. Additional details on the definition of these and other covariates can be found in the study protocol [25].

### 2.6 Statistical Analyses

We summarized patterns of vaccine uptake and described the characteristics of the Medicare FFS population who received a 2023–2024 COVID-19 vaccination. For each vaccine brand and outcome, we also summarized outcome counts and estimated the crude incidence rates of outcomes during the post-vaccination risk and control intervals.

To ensure timely safety monitoring, we conducted an early phase of SCCS analyses in the Medicare FFS database; additional details on the analysis can be found in the study protocol [25]. For the early phase analyses, SCCS analyses were conducted for adequately powered analyses for each brand and outcome, defined as those analyses with outcomes with a sufficient number of cases to detect a minimum incidence rate ratio (IRR) of at least 10 at 80% power with a two-sided alpha of 0.01. For the end-of-season analyses, we conducted SCCS analyses for any brand and outcome analyses that were powered in the early phase analysis. We also included any brand and outcome analyses that had gained a sufficient number of cases to detect a minimum IRR of at least 3 at 80% power with a two-sided alpha of 0.01. For these end-of-season brand and outcome analyses, we produced additional case-specific descriptive summaries, and conducted inferential primary, secondary, and sensitivity SCCS analyses. We present only the results of the end-of-season analyses because these results included additional data resulting in more precise estimates.

In the primary SCCS analyses, for each vaccine brand and outcome, we used conditional Poisson regression to estimate IRRs and 99% confidence intervals (CIs) which compared outcome rates in risk and control intervals. Attributable risk (AR) estimates and 99% CIs were also estimated for each outcome per 100,000 vaccine doses by estimating the number of excess cases predicted from the regression model divided by the number of eligible vaccine doses [27]. We used two-tailed hypothesis tests, with results considered statistically significant at p-value < 0.01.

Secondary analyses were performed for each vaccine brand and outcome with an elevation in incidence rates in primary analyses and a p-value < 0.1, to assess the potential effect of same-day concomitant vaccination with COVID-19 vaccination. A higher p-value threshold than the statistical significance threshold was used to determine brand and outcomes for secondary analyses because we expected subgroup analyses to have limited statistical power and intended to explore potential influences of concomitant vaccination despite this limitation. We estimated IRRs and ARs of outcomes separately among persons who received and who did not receive any of the prespecified concomitant vaccines of interest, including influenza, RSV, pneumococcal conjugate, and zoster vaccines, on the same day as their COVID-19 vaccination [28].

Both primary and secondary analyses were conducted with adjustments for event-dependent observation time (for outcomes with high case fatality rates) [29] and seasonality (all outcomes); analyses also accounted for uncertainty from outcome misclassification (where feasible). The adjustment for event-dependent observation time developed by Farrington et al. 2011 was used to adjust for curtailed observation time across analyses for outcomes with historical case fatality rates of 10 percent or higher [29]. To determine outcomes meeting this criterion, we estimated the 30-day case fatality rate of incident outcomes in the Medicare FFS population in 2021–2022 by calculating the percent of deaths in the 30 days following each outcome among all observed outcomes. We adjusted for potential bias from seasonal outcome patterns using incidence rates of outcomes estimated from the Medicare FFS population 65 years and older from corresponding calendar months in 2022–2023, which were used as the baseline incidence of outcomes in risk and control intervals. To account for uncertainty due to outcome misclassification from our claims-based outcome definitions, we performed multiple imputation analyses based on the positive predictive value (PPV) for outcomes with PPVs available (eTable 1). The PPV was defined as the proportion of true cases among cases adjudicated through medical record review and was determined from prior vaccine safety surveillance studies. The PPV-based multiple imputation method created simulated datasets where cases were sampled with probabilities equal to the PPVs. Conditional Poisson regression was repeated on the simulated datasets and IRR and AR estimates were pooled across the simulated datasets [30]. Analyses were conducted with and without adjustments for seasonality and outcome misclassification, and we included both adjustments where feasible.

We conducted two sensitivity analyses including: (i) incorporating a 14-day washout period between risk and control intervals (all outcomes except anaphylaxis) to examine potential bias from carryover effects from vaccination contributing to control interval risk; and (ii) including individuals’ full observation period length for persons that died or disenrolled (for outcomes with a historical case fatality rate of 10 percent or higher) to evaluate potential bias from a violation of the SCCS assumption that observation length is independent of events [29]. For each outcome, sensitivity analyses were conducted including all adjustments applicable to the outcome (i.e., event-dependent observation time, seasonality) and PPV-based imputation (where feasible).

All analyses were conducted using R 4.0.3 (R Foundation for Statistical Computing, Vienna, Austria) and SAS v. 9.4 (SAS Institute Inc., Cary, NC, United States).

This study was part of public health surveillance, and therefore, it was exempted from institutional review board approval and informed consent under the Common Rule.

## 3 Results

### 3.1 Descriptive results

There were 7,560,772 Medicare FFS beneficiaries aged 65 years and older who received 8,090,781 COVID-19 vaccine doses (Table 2). The majority of vaccine recipients (7,033,481; 93.03%) received only a single dose of a COVID-19 vaccine with a relatively even distribution of people who received Pfizer-BioNTech (3,689,356, 48.80%) or Moderna (3,841,245, 50.80%) COVID-19 vaccines. Very few beneficiaries received a Novavax COVID-19 vaccine (30,171, 0.40%). For Pfizer-BioNTech and Moderna, the number of vaccine doses received peaked in September and October (Figure 2). There was a small increase in the number of doses received in March 2024 after CDC recommended people 65 years and older receive an additional dose of a COVID-19 vaccine.

**Figure 2.**
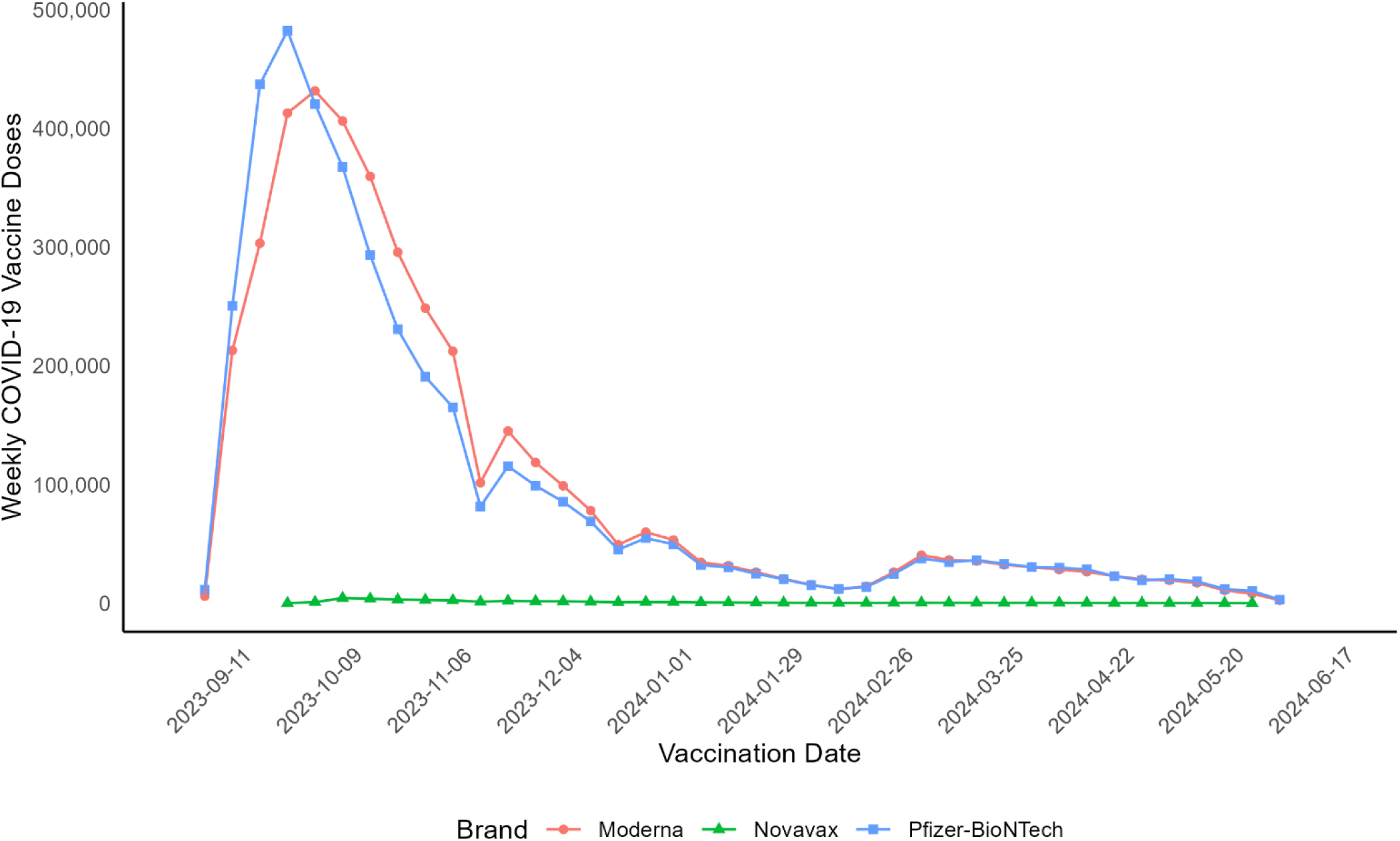
Weekly 2023–2024 COVID–19 vaccine doses by brand among Medicare Fee–for–Service beneficiaries aged ≥65 years.

**Table 2:** Characteristics of Medicare Fee–For–Service beneficiaries aged ≥65 years who received at least one dose of a 2023–2024 COVID–19 vaccine.

| Characteristic | 2023–2024 COVID–19 Vaccine Brand |  |  |  |  |  |  |  |
| --- | --- | --- | --- | --- | --- | --- | --- | --- |
|  | Any |  | Pfizer–BioNTech |  | Moderna |  | Novavax |  |
|  | N | % | N | % | N | % | N | % |
| **Total** | 7,560,772 | 100.00 | 3,689,356 | 100.00 | 3,841,245 | 100.00 | 30,171 | 100.00 |
| Age (Years) |  |  |  |  |  |  |  |  |
| 65–74 | 3,814,087 | 50.45 | 1,873,408 | 50.78 | 1,923,511 | 50.08 | 17,168 | 56.90 |
| 75–84 | 2,768,900 | 36.62 | 1,345,171 | 36.46 | 1,413,889 | 36.81 | 9,840 | 32.61 |
| ≥85 | 977,785 | 12.93 | 470,777 | 12.76 | 503,845 | 13.12 | 3,163 | 10.48 |
| Sex |  |  |  |  |  |  |  |  |
| Female | 4,290,037 | 56.74 | 2,101,168 | 56.95 | 2,171,605 | 56.53 | 17,264 | 57.22 |
| Male | 3,270,735 | 43.26 | 1,588,188 | 43.05 | 1,669,640 | 43.47 | 12,907 | 42.78 |
| Race/Ethnicity |  |  |  |  |  |  |  |  |
| Asian | 150,638 | 1.99 | 77,386 | 2.10 | 72,547 | 1.89 | 705 | 2.34 |
| Black | 353,315 | 4.67 | 180,146 | 4.88 | 172,279 | 4.48 | 890 | 2.95 |
| Hispanic | 48,576 | 0.64 | 24,731 | 0.67 | 23,700 | 0.62 | 145 | 0.48 |
| Alaskan Native/<br>American Indian | 17,334 | 0.23 | 7,264 | 0.20 | 10,035 | 0.26 | 35 | 0.12 |
| White | 6,554,473 | 86.69 | 3,184,629 | 86.32 | 3,343,467 | 87.04 | 26,377 | 87.43 |
| Other | 165,435 | 2.19 | 81,220 | 2.20 | 83,501 | 2.17 | 714 | 2.37 |
| Missing/Unknown | 271,001 | 3.58 | 133,980 | 3.63 | 135,716 | 3.53 | 1,305 | 4.33 |
| Urban/Rural |  |  |  |  |  |  |  |  |
| Urban | 6,362,709 | 84.15 | 3,232,708 | 87.62 | 3,104,647 | 80.82 | 25,354 | 84.03 |
| Rural | 1,196,312 | 15.82 | 455,711 | 12.35 | 735,797 | 19.16 | 4,804 | 15.92 |
| Missing/Unknown | 1,751 | 0.02 | 937 | 0.03 | 801 | 0.02 | 13 | 0.04 |
| Nursing Home Residency<br>Status |  |  |  |  |  |  |  |  |
| Nursing Home | 162,443 | 2.15 | 64,264 | 1.74 | 97,195 | 2.53 | 984 | 3.26 |
| Non–Nursing Home | 7,398,329 | 97.85 | 3,625,092 | 98.26 | 3,744,050 | 97.47 | 29,187 | 96.74 |
| Medicare–Medicaid Dual–<br>Eligibility |  |  |  |  |  |  |  |  |
| Dual–Eligible | 482,787 | 6.39 | 219,961 | 5.96 | 260,789 | 6.79 | 2,037 | 6.75 |
| Non–Dual–Eligible | 7,077,985 | 93.61 | 3,469,395 | 94.04 | 3,580,456 | 93.21 | 28,134 | 93.25 |
| Same–Day Concomitant<br>Vaccination |  |  |  |  |  |  |  |  |
| Any Vaccine | 3,256,914 | 43.08 | 1,647,196 | 44.65 | 1,601,607 | 41.69 | 8,111 | 26.88 |
| Influenza Vaccine | 2,764,505 | 36.56 | 1,411,565 | 38.26 | 1,346,644 | 35.06 | 6,296 | 20.87 |
| Pneumococcal Vaccine | 142,937 | 1.89 | 70,940 | 1.92 | 71,439 | 1.86 | 558 | 1.85 |
| Zoster Vaccine | 688,085 | 9.10 | 337,356 | 9.14 | 348,760 | 9.08 | 1,969 | 6.53 |
| RSV Vaccine | 95,689 | 1.27 | 47,009 | 1.28 | 48,283 | 1.26 | 307 | 1.02 |
| 2023–2024 COVID–19<br>Vaccine Dose Number |  |  |  |  |  |  |  |  |
| Dose 1 | 7,033,481 | 93.03 | 3,423,806 | 92.80 | 3,580,642 | 93.22 | 29,033 | 96.23 |
| Dose 2 | 524,964 | 6.94 | 264,228 | 7.16 | 259,622 | 6.76 | 1,114 | 3.69 |
| Dose 3 | 2,026 | 0.03 | 1,050 | 0.03 | 952 | 0.02 | 24 | 0.08 |
| Dose 4+ | 301 | 0.00 | 272 | 0.01 | 29 | 0.00 | — | — |

Acronyms: RSV: Respiratory syncytial virus

COVID-19 vaccine recipients tended to be white (86.69%), female (56.74%), of urban residency (84.15%), and non-dual-eligible for Medicare and Medicaid (93.61%) (Table 2). The majority of vaccine recipients did not reside in a nursing home (97.85%) at the time of vaccination. There was a high prevalence of people receiving concomitant vaccination across vaccine brands, and concomitant vaccination was higher among Pfizer-BioNTech (44.65%) and Moderna (41.69%) compared to Novavax (26.88%) vaccine recipients. Across brands, influenza vaccines were most frequently administered concomitantly with COVID-19 vaccines (Pfizer-BioNTech: 38.26%; Moderna: 35.06%; Novavax: 20.87%).

Outcome counts, person-time, and incidence rates for prespecified health outcomes by risk and control intervals can be found in eTable 2. The people who experienced at least one of the study outcomes during their risk or control periods tended to be slightly older (aged ≥75 years among Pfizer-BioNTech (41.51–100.00%) and Moderna (25.00–73.63%)) compared to the overall vaccinated population (Table 2, eTables 3–4). There was also a high prevalence of persons who were immunocompromised (Pfizer-BioNTech: 18.75–61.03%; Moderna: 16.67– 58.51%) and had multiple health conditions (≥2 Charlson Comorbidity Index) (Pfizer-BioNTech: 25.00–77.50%; Moderna: 31.25–77.28%). Few cases were observed across outcomes for Novavax vaccine brands because of low vaccine uptake (eTable 5).

### 3.2 Inferential results

For Pfizer-BioNTech and Moderna COVID-19 vaccine brands, the power requirements for implementing primary SCCS analyses (refer to Section 2.6) were met for all outcomes except transverse myelitis. For Novavax COVID-19 vaccines, these criteria were met for only acute myocardial infarction, deep vein thrombosis, non-hemorrhagic stroke, non-hemorrhagic stroke or TIA, pulmonary embolism and thrombosis with thrombocytopenia. However, because of the limited case count, we were unable to generate reliable estimates for non-hemorrhagic stroke and pulmonary embolism. Across the brands and outcomes analyzed, we only observed a statistically significant elevation in risk for anaphylaxis associated with Pfizer-BioNTech COVID-19 vaccines; however, this result was no longer statistically significant in analyses that included adjustment for seasonality and outcome misclassification. No conclusive evidence of an increased risk was found across vaccine brands for any other study outcomes.

We presented SCCS results from the seasonality-adjusted and the combined seasonality-adjusted and PPV-based imputation analysis for anaphylaxis. For all other outcomes, we only included results with all adjustments, but additional estimates are presented in the supplementary materials (eTables 6–19).

#### 3.2.1 Anaphylaxis

For Pfizer-BioNTech vaccine recipients, we observed an elevation in anaphylaxis risk that was statistically significant in the seasonality-adjusted primary analysis (IRR: 4.04 [99% confidence interval (CI): 1.07, 15.30]); however, it was no longer statistically significant when additionally adjusted for outcome misclassification (IRR: 3.90 [99% CI: 0.49, 30.90]) (Figure 3, eTable 6). In analyses adjusted for seasonality and PPV-based imputation, anaphylaxis cases attributable to vaccination were rare and non-statistically significant (AR per 100,000 doses: 0.09 [99% CI: −0.08, 0.25]).

**Figure 3:**
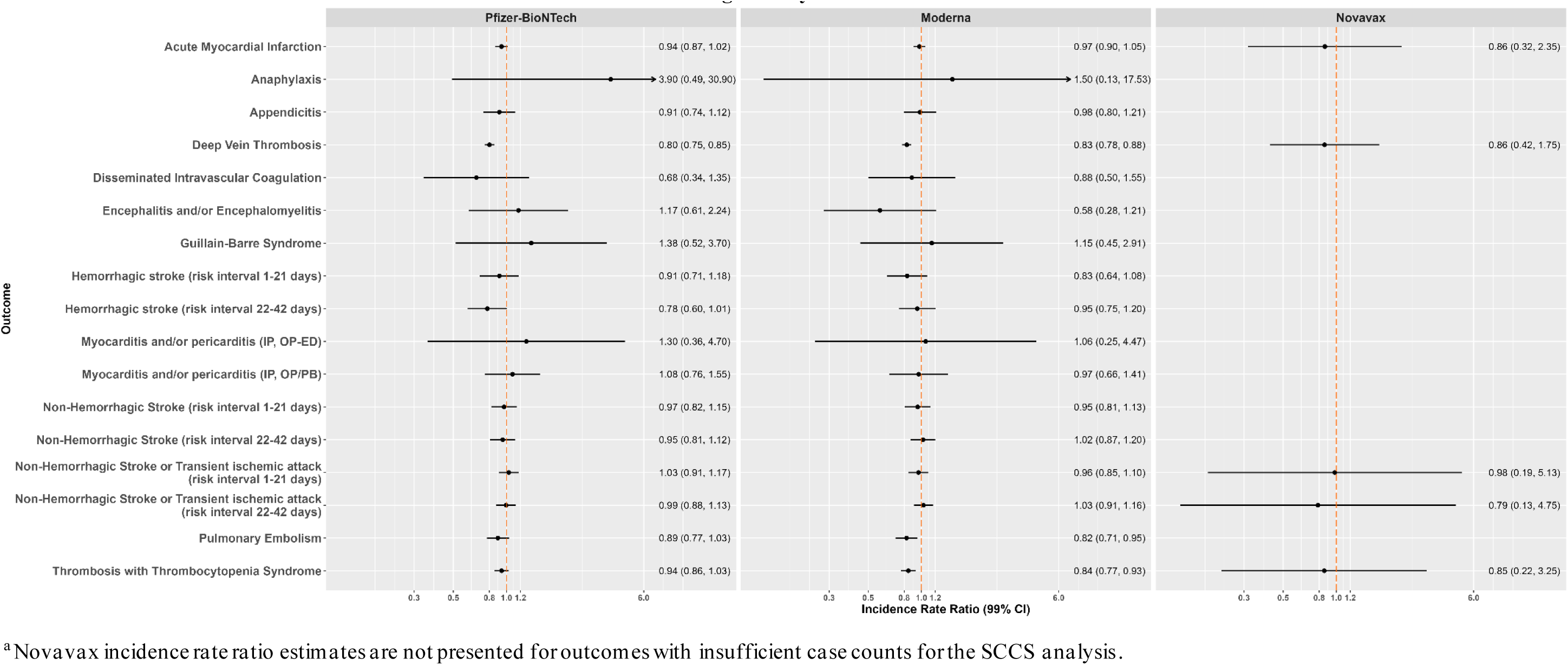
Forest plot of primary analysis outcome incidence rate ratios (IRRs) and 99% confidence intervals (CIs) by outcome and brand, for analyses including all adjustments, among Medicare Fee–for–Service beneficiaries aged ≥65 years^a^.

Secondary analyses stratifying by same-day concomitant vaccination status were only conducted for anaphylaxis following Pfizer-BioNTech vaccination. In seasonality-adjusted secondary analyses, we observed a non-statistically significant elevation in anaphylaxis risk for those with any concomitant vaccination (IRR: 2.17 [99% CI: 0.26, 17.79]), and a statistically significant elevation in risk for those without any concomitant vaccination (IRR: 6.98 [99% CI: 1.13, 43.12]) (eTable 6). When accounting for seasonality and outcome misclassification, the non-statistically significant elevation in anaphylaxis risk among those with any concomitant vaccination persisted (IRR: 2.46 [99% CI: 0.13, 45.10]). However, the increase in anaphylaxis risk among those without any concomitant vaccination was no longer statistically significant (IRR: 7.19 [99% CI: 0.43, 121.08]). In secondary analyses adjusted for seasonality and PPV-based imputation, anaphylaxis cases attributable to vaccination were rare and non-statistically significant for subgroups with any concomitant vaccination (AR per 100,000 doses: 0.06 [99% CI: −0.15, 0.27]) and without any concomitant vaccination (AR per 100,000 doses: 0.13 [99% CI: −0.09, 0.34]).

For Moderna vaccine recipients, the seasonality-adjusted primary analysis results showed a non-statistically significant elevation in anaphylaxis risk (IRR: 1.53 [99% CI: 0.29, 7.97]), which persisted in the analysis that additionally accounted for outcome misclassification (IRR: 1.50 [99% CI: 0.13, 17.53]) (Figure 3, eTable 6). Anaphylaxis cases attributable to vaccination were rare and non-statistically significant in the analysis adjusted for seasonality and PPV-based imputation (AR per 100,000 doses: 0.02 [99% CI: −0.10, 0.14]).

#### 3.2.2 Cardiovascular and Vascular Outcomes

For Pfizer-BioNTech vaccine recipients, there was no statistically significant increased risk of any cardiovascular outcomes following vaccination, including for acute myocardial infarction (IRR: 0.94 [99% CI: 0.87, 1.02]) or myocarditis/pericarditis cases in either the inpatient or outpatient emergency department (OP-ED), or in any care setting (respectively, IRR: 1.30 [99% CI: 0.36, 4.70]; IRR: 1.08 [99% CI: 0.76, 1.55]) (Figure 3; eTables 7-9). This was consistent for stroke outcomes, which included hemorrhagic stroke in the 1–21 or 22–42 day risk intervals (respectively, IRR: 0.91 [99% CI: 0.71, 1.18]; IRR: 0.78 [99% CI: 0.60, 1.01]), non-hemorrhagic stroke in the 1–21 or 22–42 day risk intervals (respectively, IRR: 0.97 [99% CI: 0.82, 1.15]; IRR: 0.95 [99% CI: 0.81, 1.12]), and non-hemorrhagic stroke or TIA in the 1–21 or 22–42 day risk intervals (respectively, IRR: 1.03 [99% CI: 0.91, 1.17]; IRR: 0.99 [99% CI: 0.88, 1.13]) (Figure 3, eTables 10–12). There was also no increased risk of vascular outcomes, which included deep vein thrombosis (IRR: 0.80 [99% CI: 0.75, 0.85]) and pulmonary embolism (IRR: 0.89 [99% CI: 0.77, 1.03]) (Figure 3, eTables 13–14).

Moderna vaccine recipients similarly showed no statistically significant elevation in risk of any cardiovascular outcomes following vaccination, including for acute myocardial infarction (IRR: 0.97 [99% CI: 0.90, 1.05]) or myocarditis/pericarditis in either the inpatient or OP-ED, or in any care setting (respectively, IRR: 1.06 [99% CI: 0.25, 4.47]; IRR: 0.97 [99% CI: 0.66, 1.41]). This was also consistent for stroke outcomes, including hemorrhagic stroke in the 1–21 or 22–42 day risk intervals (respectively, IRR: 0.83 [99% CI: 0.64, 1.08]; IRR: 0.95 [99% CI: 0.75, 1.20]), non-hemorrhagic stroke in the 1–21 or 22–42 day risk intervals (respectively, IRR: 0.95 [99% CI: 0.81, 1.13]; IRR: 1.02 [99% CI: 0.87, 1.20]), and non-hemorrhagic stroke or TIA in the 1–21 or 22–42 day risk intervals (respectively, IRR: 0.96 [99% CI: 0.85, 1.10]; IRR: 1.03 [99% CI: 0.91, 1.16]) (Figure 3, eTables 7–12). There was also no increased risk of vascular outcomes following Moderna COVID-19 vaccination, which included deep vein thrombosis (IRR: 0.83 [99% CI: 0.78, 0.88]) and pulmonary embolism (IRR: 0.82 [99% CI: 0.71, 0.95]) (Figure 3, eTables 13–14).

For Novavax vaccine recipients, there was also no increased risk of any cardiovascular outcomes following vaccination, which included acute myocardial infarction (IRR: 0.86 [99% CI: 0.32, 2.35]) and non-hemorrhagic stroke or TIA in the 1–21 or 22–42 day risk intervals (respectively, IRR: 0.98 [99% CI: 0.19, 5.13]; IRR: 0.79 [99% CI: 0.13, 4.75]) (Figure 3, eTables 7 and 12). Similarly, there was no elevated risk of deep vein thrombosis (IRR: 0.86 [99% CI: 0.42, 1.75]), which was the only vascular outcome studied (Figure 3, eTable 13). We were unable to evaluate the risk of non-hemorrhagic stroke or pulmonary embolism among Novavax recipients due to the limited number of cases (eTable 5).

#### 3.2.3 Hematological Outcomes

There was no increased risk of thrombosis with thrombocytopenia syndrome following Pfizer-BioNTech (IRR: 0.94 [99% CI: 0.86, 1.03]), Moderna (IRR: 0.84 [99% CI: 0.77, 0.93]), or Novavax (IRR: 0.85 [99% CI: 0.22, 3.25]) vaccination (Figure 3, eTable 15). Similarly, there was no elevation in risk of disseminated intravascular coagulation following Pfizer-BioNTech (IRR: 0.68 [99% CI: 0.34, 1.35]) or Moderna (IRR: 0.88 [99% CI: 0.50, 1.55]) vaccination (Figure 3, eTable 16).

#### 3.2.4 Neurologic Outcomes

There was no association between Pfizer-BioNTech COVID-19 vaccination and GBS (IRR: 1.38 [99% CI: 0.52, 3.70]) or encephalitis/encephalomyelitis (IRR: 1.17 [99% CI: 0.61, 2.24]) (Figure 3, eTables 17–18). Similarly, there was no association between Moderna COVID-19 vaccination and GBS (IRR: 1.15 [99% CI: 0.45, 2.91)) or encephalitis/encephalomyelitis (IRR: 0.58 [99% CI: 0.28, 1.21]).

#### 3.2.5 Other Outcomes

There was no association between appendicitis and Pfizer-BioNTech (IRR: 0.91 [99% CI: 0.74, 1.12]) or Moderna (IRR: 0.98 [99% CI: 0.80, 1.21]) COVID-19 vaccination (Figure 3, eTable 19).

### 3.3 Sensitivity analyses

Results of the sensitivity analyses incorporating a 14-day washout period between risk and control intervals were similar to the primary analyses (eTables 6–19). Results were also consistent when we included an individuals’ full observation period length for persons that died or disenrolled for outcomes with a historical case fatality rate of 10 percent or higher.

## 4 Discussion

Our study evaluated the association between COVID-19 vaccination and multiple health outcomes among persons 65 years and older in one of the largest medical health plan databases in the United States. A rare but potential elevation in anaphylaxis risk was observed following Pfizer-BioNTech vaccination that was no longer statistically significant in analyses accounting for seasonality and outcome misclassification. No statistically significant elevations in risk were observed for other outcomes studied following Pfizer-BioNTech, Moderna or Novavax COVID-19 vaccination.

This study’s potential, but rare, elevation in risk for anaphylaxis observed following COVID-19 vaccination among persons 65 years and older is consistent with findings from existing safety assessments performed in the U.S. and internationally across various COVID-19 vaccine formulations and seasons [31–33]. A study of U.S and European passive surveillance systems that included data from December 2020 through mid-August 2021 estimated the mean anaphylaxis rate following COVID-19 vaccination was 10.67 cases per million doses, which was comparable to the herpes zoster vaccine [33]. However, various reports have demonstrated that post-vaccination incidence rates of anaphylaxis have decreased since the authorization of COVID-19 vaccines [31, 32]. In our study, the estimated excess rate of anaphylaxis cases following 2023–2024 Pfizer-BioNTech COVID-19 vaccination based on the self-controlled model was less than 1 case per million doses. Estimates from subgroup analyses for those with and without a same-day concomitant vaccination were very imprecise and overlapped. Based on the data, there was insufficient evidence to conclude risk increased with same-day concomitant vaccination.

Our study did not find evidence for a statistically significant elevation in hemorrhagic or non-hemorrhagic stroke risk associated with COVID-19 vaccination among Medicare beneficiaries aged 65 years and older. This is largely consistent with the available literature from safety assessments of bivalent COVID-19 mRNA vaccines [14–16, 18, 19, 34]. Several studies evaluating the safety of bivalent COVID-19 mRNA vaccines have found no elevation in hemorrhagic stroke risk following vaccine administration [14, 16, 34]. Multiple studies have also shown no conclusive elevation in the risk of non-hemorrhagic stroke associated with these vaccines administered alone [14–16, 18, 19, 34]. However, a few studies have suggested a small but potential increase in the risk of non-hemorrhagic stroke associated with concomitant bivalent COVID-19 mRNA and high-dose or adjuvanted influenza vaccination [14, 35]. Similarly, the Vaccine Safety Datalink (VSD) also detected a statistical signal for ischemic stroke (i.e., non-hemorrhagic stroke) in the 1–42 days following 2023–2024 Moderna COVID-19 vaccination in the population 65 years and older [36]. However, results were inconsistent across age groups and risk intervals, and the available data did not support clear and consistent evidence of any increased risk of ischemic stroke. This conclusion is consistent with our study results; we did not find evidence for an elevation in non-hemorrhagic stroke risk following vaccination with any of the vaccine brands, with no elevations in risk observed for several analyses. Where elevations in risk were observed, these were small and non-statistically significant.

In our study, we did not find any associations between COVID-19 vaccine brands and GBS. This differed from VSD where there was a statistical signal for GBS in the 1–42 days following 2023–2024 Pfizer-BioNTech COVID-19 vaccination in the population 65 years and older [36]. However, VSD and FDA have not detected any associations between any COVID-19 vaccine brand and GBS in any prior studies [9, 36, 37]. Based on our data, there is insufficient evidence to conclude that there is any increased risk of GBS following 2023–2024 Pfizer-BioNTech COVID-19 vaccination.

Other study findings remained consistent with safety assessments from prior COVID-19 vaccine formulations conducted in the same database and age group. No elevation in risk was observed for acute myocardial infarction [38]. We similarly found no statistically significant elevation in the risk of myocarditis/pericarditis following COVID-19 vaccination [38]. Prior studies conducted found inconsistent evidence related to pulmonary embolism risk following COVID-19 vaccine administration; primary series vaccines were associated with a statistically significant increase in risk, whereas monovalent booster vaccines were associated with a reduction in risk [38]. This study observed no elevation in pulmonary embolism risk following COVID-19 vaccines, consistent with findings following the booster vaccine [38].

The study has several strengths. It leveraged health data from one of the largest medical insurance claims databases for individuals 65 years and older in the United States. This improved the generalizability of the results and allowed for the evaluation of elevations in risk for particularly rare outcomes. The SCCS study design which compares risk within individuals also allowed for the control of time invariant person-level risk factors that could bias the estimation of risk for outcomes (i.e., sex, the presence of chronic conditions, etc.). To assess the robustness of study results, our study adjusted for seasonality in estimating the post-vaccination risk of outcomes. Where feasible, we also accounted for uncertainty in the accuracy of claims-based outcome definitions using PPV-based imputation that simulated the analysis using cases sampled based on the proportion of chart-confirmed cases estimated from prior safety assessments in the same database and age group.

There were several limitations to the study. Our study used claims-based outcome definitions that are subject to outcome misclassification. While PPV-based imputation helps to account for uncertainty related to outcome misclassification, PPV-based imputation does not fully control for outcome misclassification as accurately as restricting the analysis to within-study chart-confirmed cases. Outcome misclassification could therefore still be present and bias risk estimates in either direction. Additionally, the SCCS design requires clearly defined risk intervals to estimate risk attributed to vaccination. For outcomes such as stroke with varied evidence on their respective periods of vaccine-attributable risk following vaccination, we used multiple risk interval definitions. Where feasible, we also conducted sensitivity analyses including a 14-day washout period. Despite these measures, and our use of risk intervals determined based on literature and clinician input, the possibility for misspecification of risk intervals still exists for these and other study outcomes and could result in misattributed risk to vaccines. Lastly, the study may be subject to time-varying confounders that are not adjusted for in the model. These factors could bias risk estimates in either direction.

This study found a small but potential elevation in anaphylaxis risk associated with the 2023–2024 Pfizer-BioNTech COVID-19 vaccines, and no evidence for an increased risk of other outcomes following vaccination with 2023–2024 COVID-19 vaccines. Our study contributes to growing evidence on the safety of COVID-19 vaccines. FDA continues to believe that the benefits of COVID-19 vaccination outweigh the risks.

## Supporting information

Supplementary_File_2023_24_COVID-19_Vaccine_Surveillance_01032025.xlsx

## Data Availability

All data produced in the present study are available upon reasonable request to the authors.

## Acknowledgments

We would like to thank Yue Wu (Acumen LLC), Bradley Lufkin (Acumen LLC), Bowen Chen (Acumen LLC), Kira Lin (Acumen LLC), Nimesh Shah (Acumen LLC), Purva Shah (Acumen LLC), Samikshya Siwakoti (Acumen LLC), Godwin Anguzu (Acumen LLC), Merianne Spencer (FDA), and Jane Gwira (FDA).

## References

1. FDA Approves First COVID-19 Vaccine [press release]. 2021.[accessed October, 2023]. Available from: https://www.fda.gov/news-events/press-announcements/fda-approves-first-covid-19-vaccine.

2. Ikeokwu AE, Lawrence R, Osieme ED, Gidado KM, Guy C, Dolapo O. Unveiling the Impact of COVID-19 Vaccines: A Meta-Analysis of Survival Rates Among Patients in the United States Based on Vaccination Status. Cureus. 2023;15(8):e43282.

3. Layton JB, Peetluk L, Wong HL, Jiao Y, Djibo DA, Bui C, et al. Effectiveness of monovalent COVID-19 booster/additional vaccine doses in the United States. Vaccine: X. 2024;16:100447.

4. Lu Y, Lindaas A, Matuska K, Izurieta HS, McEvoy R, Menis M, et al. Real-world Effectiveness of mRNA COVID-19 Vaccines Among US Nursing Home Residents Aged ≥65 Years in the Pre-Delta and High Delta Periods. Open forum infectious diseases. 2024;11(3):ofae051.

5. Centers for Disease Control and Prevention. Adults 65+ Coverage: Updated 2023-24 COVID-19 Vaccination Coverage, Adults 65 Years and Older, United States 2023.[accessed October, 2023]. Available from: https://www.cdc.gov/vaccines/imz-managers/coverage/covidvaxview/interactive/adults-65yrs-over-coverage.html.

6. Farshbafnadi M, Kamali Zonouzi S, Sabahi M, Dolatshahi M, Aarabi MH. Aging & COVID-19 susceptibility, disease severity, and clinical outcomes: The role of entangled risk factors. Experimental gerontology. 2021;154:111507.

7. Centers for Disease Control and Prevention. Underlying Medical Conditions Associated with Higher Risk for Severe COVID-19: Information for Healthcare Professionals 2024.[accessed October, 2023]. Available from: https://www.cdc.gov/covid/hcp/clinical-care/underlying-conditions.html.

8. Shoaibi A, Lloyd PC, Wong H-L, Clarke TC, Chillarige Y, Do R, et al. Evaluation of potential adverse events following COVID-19 mRNA vaccination among adults aged 65 years and older: Two self-controlled studies in the U.S. Vaccine. 2023;41(32):4666–78.

9. Wong HL, Tworkoski E, Ke Zhou C, Hu M, Thompson D, Lufkin B, et al. Surveillance of COVID-19 vaccine safety among elderly persons aged 65 years and older. Vaccine. 2023;41(2):532–9.

10. Jabagi MJ, Botton J, Bertrand M, Weill A, Farrington P, Zureik M, et al. Myocardial Infarction, Stroke, and Pulmonary Embolism After BNT162b2 mRNA COVID-19 Vaccine in People Aged 75 Years or Older. Jama. 2022;327(1):80–2.

11. Takeuchi Y, Iwagami M, Ono S, Michihata N, Uemura K, Yasunaga H. A post-marketing safety assessment of COVID-19 mRNA vaccination for serious adverse outcomes using administrative claims data linked with vaccination registry in a city of Japan. Vaccine. 2022;40(52):7622–30.

12. Whiteley WN, Ip S, Cooper JA, Bolton T, Keene S, Walker V, et al. Association of COVID-19 vaccines ChAdOx1 and BNT162b2 with major venous, arterial, or thrombocytopenic events: A population-based cohort study of 46 million adults in England. PLoS medicine. 2022;19(2):e1003926.

13. Shimabukuro T, editor Update on COVID-19 and influenza vaccine safety. Advisory Committee on Immunization Practices (ACIP); 2024: Centers for Disease Control and Prevention,.

14. Lu Y, Matuska K, Nadimpalli G, Ma Y, Duma N, Zhang HT, et al. Stroke Risk After COVID-19 Bivalent Vaccination Among US Older Adults. Jama. 2024;331(11):938–50.

15. Yamin D, Yechezkel M, Arbel R, Beckenstein T, Sergienko R, Duskin-Bitan H, et al. Safety of monovalent and bivalent BNT162b2 mRNA COVID-19 vaccine boosters in at-risk populations in Israel: a large-scale, retrospective, self-controlled case series study. The Lancet Infectious Diseases. 2023;23(10):1130–42.

16. Andrews N, Stowe J, Miller E, Ramsay M. BA.1 Bivalent COVID-19 Vaccine Use and Stroke in England. Jama. 2023;330(2):184–5.

17. Jabagi M-J, Bertrand M, Botton J, Vu SL, Weill A, Dray-Spira R, et al. Stroke, Myocardial Infarction, and Pulmonary Embolism after Bivalent Booster. 2023;388(15):1431–2.

18. Gorenflo MP, Davis PB, Kaelber DC, Xu R. Ischemic stroke after COVID-19 bivalent vaccine administration in patients aged 65 years and older in the United States. NPJ vaccines. 2023;8(1):180.

19. Xu S, Sy LS, Hong V, Holmquist KJ, Qian L, Farrington P, et al. Ischemic Stroke after Bivalent COVID-19 Vaccination: A Self-Controlled Case Series Study. 2023:2023.10.12.23296968.

20. FDA Takes Action on Updated mRNA COVID-19 Vaccines to Better Protect Against Currently Circulating Variants [press release]. 2023.[accessed October, 2023]. Available from: https://www.fda.gov/news-events/press-announcements/fda-takes-action-updated-mrna-covid-19-vaccines-better-protect-against-currently-circulating.

21. United States Food and Drug Administration. Novavax COVID-19 Vaccine, Adjuvanted (2023-2024) Letter of Authorization. 2023.

22. Older Adults Now Able to Receive Additional Dose of Updated COVID-19 Vaccine [press release]. 2024.[accessed October, 2024]. Available from: https://www.cdc.gov/media/releases/2024/s-0228-covid.html.

23. Centers for Disease Control and Prevention. Use of Updated COVID-19 Vaccines 2023–2024 Formula for Persons Aged ≥6 Months: Recommendations of the Advisory Committee on Immunization Practices — United States, September 2023. 2023 October, 2023.

24. Hallas J, Pottegård A. Use of self-controlled designs in pharmacoepidemiology. Journal of internal medicine. 2014;275(6):581–9.

25. Gruber FJ, Zelaya EC, Forshee R, Anderson AS, Ondari M, Hu M, et al. Evaluation of Multiple Safety Outcomes following 2023-2024 COVID-19 Vaccination in Persons 6 Months and Older. In: Department of Health and Human Services, editor. 2023.

26. Centers for Disease Control and Prevention. Interim Clinical Considerations for Use of COVID-19 Vaccines in the United States 2023.[accessed October, 2023]. Available from: https://www.cdc.gov/vaccines/covid-19/clinical-considerations/interim-considerations-us.html.

27. Cox C, Li X. Model-Based Estimation of the Attributable Risk: A Loglinear Approach. Computational statistics & data analysis. 2012;56(12):4180–9.

28. Centers for Disease Control and Prevention. Recommended Vaccines by Age 2023. Available from: https://www.cdc.gov/vaccines/vpd/vaccines-age.html.

29. Farrington CP, Anaya-Izquierdo K, Whitaker HJ, Hocine MN, Douglas I, Smeeth L. Self-Controlled Case Series Analysis With Event-Dependent Observation Periods. Journal of the American Statistical Association. 2011;106(494):417–26.

30. Rubin DB, Schenker N. Multiple Imputation for Interval Estimation From Simple Random Samples With Ignorable Nonresponse. Journal of the American Statistical Association. 1986;81(394):366–74.

31. Jaggers J, Wolfson AR. mRNA COVID-19 Vaccine Anaphylaxis: Epidemiology, Risk Factors, and Evaluation. Current allergy and asthma reports. 2023;23(3):195–200.

32. Boufidou F, Hatziantoniou S, Theodoridou K, Maltezou HC, Vasileiou K, Anastassopoulou C, et al. Anaphylactic Reactions to COVID-19 Vaccines: An Updated Assessment Based on Pharmacovigilance Data. Vaccines. 2023;11(3).

33. Maltezou HC, Anastassopoulou C, Hatziantoniou S, Poland GA, Tsakris A. Anaphylaxis rates associated with COVID-19 vaccines are comparable to those of other vaccines. Vaccine. 2022;40(2):183–6.

34. Jabagi MJ, Bertrand M, Botton J, Le Vu S, Weill A, Dray-Spira R, et al. Stroke, Myocardial Infarction, and Pulmonary Embolism after Bivalent Booster. The New England journal of medicine. 2023;388(15):1431–2.

35. Panagiotakopoulos LG, Monica; Moulia, L. Danielle; Link-Gelles, Ruth; Taylor, A. Christopher; Chatham-Stephens, Kevin; Brooks, Oliver; Daley, F. Matthew; Fleming, E. Katherine-Dutra; Wallace, Megan. Use of an Additional Updated 2023–2024 COVID-19 Vaccine Dose for Adults Aged ≥65 Years: Recommendations of the Advisory Committee on Immunization Practices — United States, 2024. Centers for Disease Control and Prevention,; 2024.

36. Duffy J. COVID-19 vaccine safety surveillance for the 2023-2024 season. Presentation. Office IS; 2024.

37. Lloyd PC, Smith ER, Gruber JF, Ondari M, Wong HL, Hu M, et al. Safety Monitoring of Bivalent COVID-19 mRNA Vaccines Among Recipients 6 months and Older in the United States. 2024:2024.01.24.24301676.

38. Shoaibi A, Lloyd PC, Wong HL, Clarke TC, Chillarige Y, Do R, et al. Evaluation of potential adverse events following COVID-19 mRNA vaccination among adults aged 65 years and older: Two self-controlled studies in the U.S. Vaccine. 2023;41(32):4666–78.

39. Smeeth L, Thomas SL, Hall AJ, Hubbard R, Farrington P, Vallance P. Risk of myocardial infarction and stroke after acute infection or vaccination. The New England journal of medicine. 2004;351(25):2611–8.

40. Su JR, Moro PL, Ng CS, Lewis PW, Said MA, Cano MV. Anaphylaxis after vaccination reported to the Vaccine Adverse Event Reporting System, 1990-2016. The Journal of allergy and clinical immunology. 2019;143(4):1465–73.

41. Donahue JG, Kieke BA, Lewis EM, Weintraub ES, Hanson KE, McClure DL, et al. Near Real-Time Surveillance to Assess the Safety of the 9-Valent Human Papillomavirus Vaccine. Pediatrics. 2019;144(6).

42. Kearon C, Akl EA. Duration of anticoagulant therapy for deep vein thrombosis and pulmonary embolism. Blood. 2014;123(12):1794–801.

43. Vickers ER, McClure DL, Naleway AL, Jacobsen SJ, Klein NP, Glanz JM, et al. Risk of venous thromboembolism following influenza vaccination in adults aged 50years and older in the Vaccine Safety Datalink. Vaccine. 2017;35(43):5872–7.

44. Tang N, Li D, Wang X, Sun Z. Abnormal coagulation parameters are associated with poor prognosis in patients with novel coronavirus pneumonia. Journal of thrombosis and haemostasis : JTH. 2020;18(4):844–7.

45. Pellegrino P, Carnovale C, Perrone V, Pozzi M, Antoniazzi S, Clementi E, et al. Acute disseminated encephalomyelitis onset: evaluation based on vaccine adverse events reporting systems. PloS one. 2013;8(10):e77766.

46. Schonberger LB, Bregman DJ, Sullivan-Bolyai JZ, Keenlyside RA, Ziegler DW, Retailliau HF, et al. Guillain-Barre syndrome following vaccination in the National Influenza Immunization Program, United States, 1976--1977. American journal of epidemiology. 1979;110(2):105–23.

47. Shimabukuro T, editor mRNA COVID-19 bivalent booster vaccine safety update. Advisory Committee on Immunization Practices (ACIP); 2023.

48. Oster ME, Shay DK, Su JR, Gee J, Creech CB, Broder KR, et al. Myocarditis Cases Reported After mRNA-Based COVID-19 Vaccination in the US From December 2020 to August 2021. Jama. 2022;327(4):331–40.

49. Agmon-Levin N, Kivity S, Szyper-Kravitz M, Shoenfeld Y. Transverse myelitis and vaccines: a multi-analysis. Lupus. 2009;18(13):1198–204.

50. Hosseini R, Askari N. A review of neurological side effects of COVID-19 vaccination. European journal of medical research. 2023;28(1):102.

